# Screening for attention-deficit/hyperactivity disorder traits among adults living with HIV or taking pre-exposure prophylaxis in Switzerland: a cross-sectional survey nested within the Swiss HIV Cohort Study and SwissPrEPared - a study protocol

**DOI:** 10.64898/2026.09.08.26362490

**Authors:** David Jackson-Perry, Stephanie Campos Ochoa, Salvatore Corbisiero, Irene A. Abela, Grace H. Yoon, Matthias Cavassini, Benjamin Hampel, Alain Amstutz, the Swiss HIV Cohort Study, the SwissPrEPared Cohort Study

## Abstract

**Introduction:** People living with HIV experience disproportionately high rates of mental health conditions, yet adult attention-deficit/hyperactivity disorder (ADHD) is largely absent from the HIV literature. Traits associated with ADHD, together with social exclusion, adverse life events, stigma, and barriers to accessing healthcare may increase vulnerability to HIV acquisition and complicate adherence to treatment and prevention, and a comparable pathway may apply to users of HIV pre-exposure prophylaxis (PrEP). Prevalence data on ADHD in people living with HIV are limited to two small-scale studies, and no data exist for PrEP users. The objective of this study is to estimate the prevalence of ADHD traits among people living with HIV and PrEP users in Switzerland and to describe sociodemographic, behavioural and clinical patterns. The study was co-created and is co-led by people who, beyond their academic and clinical practices, have experiential knowledge of the topics being explored

**Methods and analysis:** This is a multicentre cross-sectional survey using a Critical ADHD Studies framework and nested within the Swiss HIV Cohort Study (SHCS) and SwissPrEPared cohort. Over a 12-month recruitment period, participants attending routine visits at participating sites will complete the six-item Adult ADHD Self-Report Scale Screener (ASRS-5). A positive screen is defined as a weighted score of 14 or higher. Three additional items capture the presence of traits in childhood before the age of 12, any previously established ADHD diagnosis, and the participant’s wish to receive their score and/or a follow-up consultation with a psychologist or psychiatrist. Screening data will be linked to routinely collected cohort variables. The primary objective is to estimate the prevalence of a positive ASRS-5 screen, with 95% Wilson score confidence intervals, in each cohort. Secondary objectives include comparing cohorts and examining associations between ADHD traits and substance use, sexual behaviour, sexually transmitted infections, depression and anxiety, treatment and prevention adherence, use of and interest in long-acting HIV treatment modalities, and gender identity and sexual orientation. A target sample of 800 participants per cohort was calculated for a 3% prevalence margin of error.

**Ethics and dissemination:** The study is nested within the SHCS and SwissPrEPared and falls under their existing ethical approvals and general participant consent. Participants who screen positive and wish to be followed up will be offered a consultation with an ADHD-experienced psychologist or psychiatrist. Results will be publicly available on the study website, published in peer-reviewed journals, and shared with participants and communities through lay summaries and cohort communication channels.

**STRENGTHS AND LIMITATIONS OF THIS STUDY:**

⍰ The first study to systematically estimate the prevalence of ADHD traits at scale among people living with HIV and among users of HIV pre-exposure prophylaxis (PrEP).
⍰ Systematic administration to all participants attending routine visits, embedded in existing cohort workflows, limiting self-selection and reducing participation bias.
⍰ Linkage to rich, routinely collected cohort data allowing exploration of a broad range of sociodemographic, behavioural, and clinical correlates at scale.
⍰ The ASRS-5 identifies ADHD traits rather than making a clinical diagnosis; the study therefore estimates screening-positive trait prevalence, expected to exceed the possible prevalence of ADHD diagnoses, as in the general population.
⍰ Differences in questionnaire administration between cohorts (provider-administered in the SHCS versus self-administered in SwissPrEPared) and incomplete harmonisation of some variables may limit between-cohort comparability, resulting in cohort-stratified analyses instead.

## INTRODUCTION

People living with HIV experience disproportionately high rates of mental health challenges, including depression, anxiety, and substance use disorders, which can undermine treatment adherence, viral suppression, and quality of life.^3,4^ However, adult attention-deficit/hyperactivity disorder (ADHD) remains largely absent from the HIV literature, and an urgent need for data at this intersection has been noted.^5^

ADHD is diagnostically characterised as a neurodevelopmental disorder affecting three core areas, inattention, impulsivity, and hyperactivity. The prevalence of ADHD amongst adults (meeting full diagnostic criteria including onset in childhood) is approximately 2.5%.^6^ However, the true prevalence is likely higher, particularly in key sub-populations such as women and racially minoritised individuals, who face increased barriers to diagnosis.^6,7^

Adults with ADHD - particularly if unrecognised or untreated - may face a range of adverse health outcomes such as co-occurring psychiatric conditions, obesity, smoking, problematic substance use, poorer sexual health, and difficulty maintaining adherence to prescribed treatment regimens, indicating the potential usefulness of long-acting HIV treatment options.^8–10^ Compared to the general population, adults with ADHD are twice as likely to die prematurely with a significantly reduced life expectancy.^11,12^ Treatment - which requires diagnosis first - improves many of these outcomes.^13,14^ However, systemic barriers to diagnosis, accessing healthcare, and social exclusion, adverse life experiences, and stigma act as aggravating factors^1,2,15^: certain negative outcomes may therefore be shaped as much by unaccommodating or hostile environments as by the condition itself. Further, due to limited professional understanding of adult ADHD, there is evidence that a high proportion of people being treated for other mental health conditions live with undiagnosed ADHD.^16^ It is imperative that clinicians be aware of ADHD to consider the possibility of co-occurrence and avoid misattribution of over-lapping traits to other conditions, thereby missing out on the benefits of diagnosis and treatment of ADHD.^17^

Converging lines of evidence suggest that a combination of social factors and traits associated with ADHD could heighten vulnerability to acquiring HIV.^5,18^ ADHD - particularly its hyperactive-impulsive and combined presentations - is associated with differences in self-regulation, reduced delay of gratification, and altered reward processing.^19,20^ These features may bias decision-making toward immediate rewards and away from longer-term consequences, complicating behaviours such as consistent condom use or delaying penetration until protection is available.^19^ Adults with ADHD also more frequently report adverse childhood experiences, victimisation, relationship instability, substance use (including in sexualised settings)^21^ and socioeconomic stress, all factors that may compound impulsivity, reduce access to protective resources and increase exposure to higher-risk sexual contexts. Further, adults with ADHD are considerably more likely to identify with sexual and/or gender minorities^22^, and to have had more same-gendered partners, irrespective of sexual identity.^23,24^ The only two small-scale studies exploring HIV and adult ADHD co-prevalence (one in India and one in Turkey) diagnosed 20-29% of adults living with HIV with ADHD, significantly above general population estimates.^25,26^

Among PrEP users, no co-prevalence data exist. PrEP users, especially gay, bisexual and other men who have sex with men, are more likely than non-PrEP users to report condomless sex, higher numbers of partners and the use of sexually enhancing substances.^27^ In a Belgian screening study, PrEP users scored four times higher than the general population for autistic traits. People living with HIV scored twice as high. With growing recognition that autism and ADHD frequently co-occur^28^, we may find a similar pattern for ADHD.

HIV, ADHD, and PrEP each carry stigma, and the intersection is complex. We therefore sought a theoretical framework that minimises the risks of exacerbating stigma while offering a more holistic understanding of these intersections, and adopted Critical ADHD Studies (CADS).^29,30^ CADS is a recent ADHD-specific offshoot of neurodiversity approaches, which hold that neurological variation is naturally occurring rather than inherently disordered, and locate disability to varying degrees in inadequate structural support (the social model of disability) rather than as purely individual pathology (the medical model). Neurodiversity approaches are, however, frequently mischaracterized as focusing solely on social constraints,^31^ and remain largely centred on autism, reflecting its roots in 1990 autistic activism.^32^ CADS retains core principles of neurodiversity approaches while offering more clearly delineated foundational precepts, and addressing ADHD specificities such as the role of medication. Four tenets are central. First, ADHDers, both academic and lay, should be central to ADHD knowledge creation: no perspective is complete, but without lived experience, knowledge remains partial. Second, CADS is founded on interdisciplinarity. While ADHD medication’s often transformational role arguably strengthens the medical model’s claims to knowledge production in the field, CADS engages the medical model alongside social sciences and humanities, attending throughout to systemic and health inequities while still recognising difficulties associated with impairment. Third, ADHD is neither a superpower nor disorder - both framings risk harms, either understating difficulties or stigmatizing those so labelled - but is instead complex, contextual, and relational, including both inherent impairments and disabling social norms and expectations. Fourth, because ADHD diagnosis and treatment access are markedly inequitable^6,7,33^, intersectionality - which explicates how social identities (race, gender, class, sexuality) overlap to produce distinct forms of discrimination, privilege, and systemic oppression - is central to CADS.

## STUDY AIMS

The primary aims are to estimate the prevalence of ADHD traits among people living with HIV and those taking PrEP and in doing so raise awareness and knowledge about ADHD among HIV healthcare providers, people living with HIV, and PrEP users. Secondary objectives are (1) to describe and compare the sociodemographic and clinical characteristics of participants with and without a positive screen; (2) to assess associations between ADHD traits and (a) substance use, sexual behaviour and sexually transmitted infections (STIs), (b) depression and anxiety, (c) adherence to treatment and/or prevention, and (d) use of and interest in long-acting HIV treatment options; (3) to describe and compare gender identity and sexual orientation among PrEP users with and without ADHD traits; and (4) to assess whether participants with ADHD traits subsequently receive a formal diagnosis.

## METHODS AND ANALYSIS

### Study design

This is a multicentre cross-sectional survey nested within two established Swiss cohorts, the Swiss HIV Cohort Study (SHCS)^34^ and SwissPrEPared, with recruitment of individual participants over a 12-month period. The design draws on and adapts a recent similar screening study for autistic traits in People living with HIV and PrEP users in Belgian sexual health clinics.^35^ Anticipated recruitment begins in November 2026.

The SHCS is an ongoing, nationwide, multicenter cohort study established in 1988 enrolling adults living with HIV in Switzerland. The cohort is estimated to include approximately 62.4% of all people diagnosed with HIV in the country.^34,36^ Participants attend study visits every six months, during which comprehensive clinical, behavioral, and laboratory data are collected during the medical visit. Since 2024, all new enrollments are asked about the reasons why they decided against the use of PrEP.

SwissPrEPared is an ongoing, nationwide, multicenter cohort study established in 2019 enrolling individuals interested in using PrEP for HIV prevention.^38^ As of 2025, 91 recruiting centers participate, located in 18 of Switzerland’s 26 cantons. Centers include tertiary and cantonal hospitals, community-based voluntary counselling and testing services (“Checkpoints”), and private medical practices, such as general practitioners, infectious diseases specialists, and dermatologists. Study visits include PrEP counselling and a medical assessment and occur at regular intervals: every three months for participants using daily PrEP and at least every six months for those using intermittent PrEP.

### Study sites

The study will initially be conducted at three sites that participate in both cohorts: Checkpoint Zurich, Lausanne University Hospital (CHUV) and University Hospital Zurich (USZ). These sites were selected because they can ensure referral and follow-up for participants who request it, are large enough to reach the target sample size, and offer some regional diversity (German and French parts of Switzerland). Other sites, including teaching hospitals and private practices, may be included depending on their interest and capacity.

### Participants

Eligible participants are people aged 18 years or older, enrolled in the SHCS or SwissPrEPared who are able to read and understand the language in which the questionnaire is administered.

### Patient and public involvement and engagement

This project is community-driven, and the research team includes members in project lead positions with experiential knowledge relevant to the research questions. This has shaped the choice of the CADS framework, and methods that better accommodate ADHD- and HIV-related characteristics that might otherwise affect data collection. In addition, the website, study tools, and publications will be reviewed by at least two external non-academic community members (one PrEP user, one person living with HIV) as well as by routine cohort physicians for relevance and clarity. Involvement and engagement will be reported using the GRIPP2 checklist^37^. Results will be made available as lay summaries through community channels, on the SHCS, SwissPrEPared, and study websites^39^ and feature in an SHCS podcast.

### Sampling and procedure

All participants attending a routine cohort visit during the 12-month study recruitment period will be invited to take part. The SHCS conducts twice-yearly visits and SwissPrEPared every three to six months, hence, a 12-month window should capture at least one visit per active participant at each site. Offering the survey systematically to all attendees, embedded in the routine visit, should yield a more unbiased sampling approach.

In the SHCS, the questionnaire will be administered by the treating physician during the routine consultation and entered into a REDCap form linked with the cohort data collection system. In SwissPrEPared, the questionnaire will be self-administered before the visit through the smartphone-based cohort application, with the possibility to fill it in during the visit for participants who prefer this. These procedures represent routine data collection processes in each cohort and thus present an optimal embedded study procedure to avoid additional burden for both providers and participants. In both cohorts a printed information leaflet will be provided. Site staff and the leaflet clarify that participation is useful even for people who are confident they do not have ADHD traits, or on the contrary have a pre-existing ADHD diagnosis, and that an elevated screening score does not constitute diagnosis. All Participants will be able to discuss their results with their physician, and those who screen positive (score ≥ 14) and express interest in a follow-up will receive timely referral for a specialist psychiatric consultation (see below). Participation is voluntary, and completing the questionnaire takes approximately 5-10 minutes.

The entire survey procedure is outlined in Figure 1.

**Figure 1.**
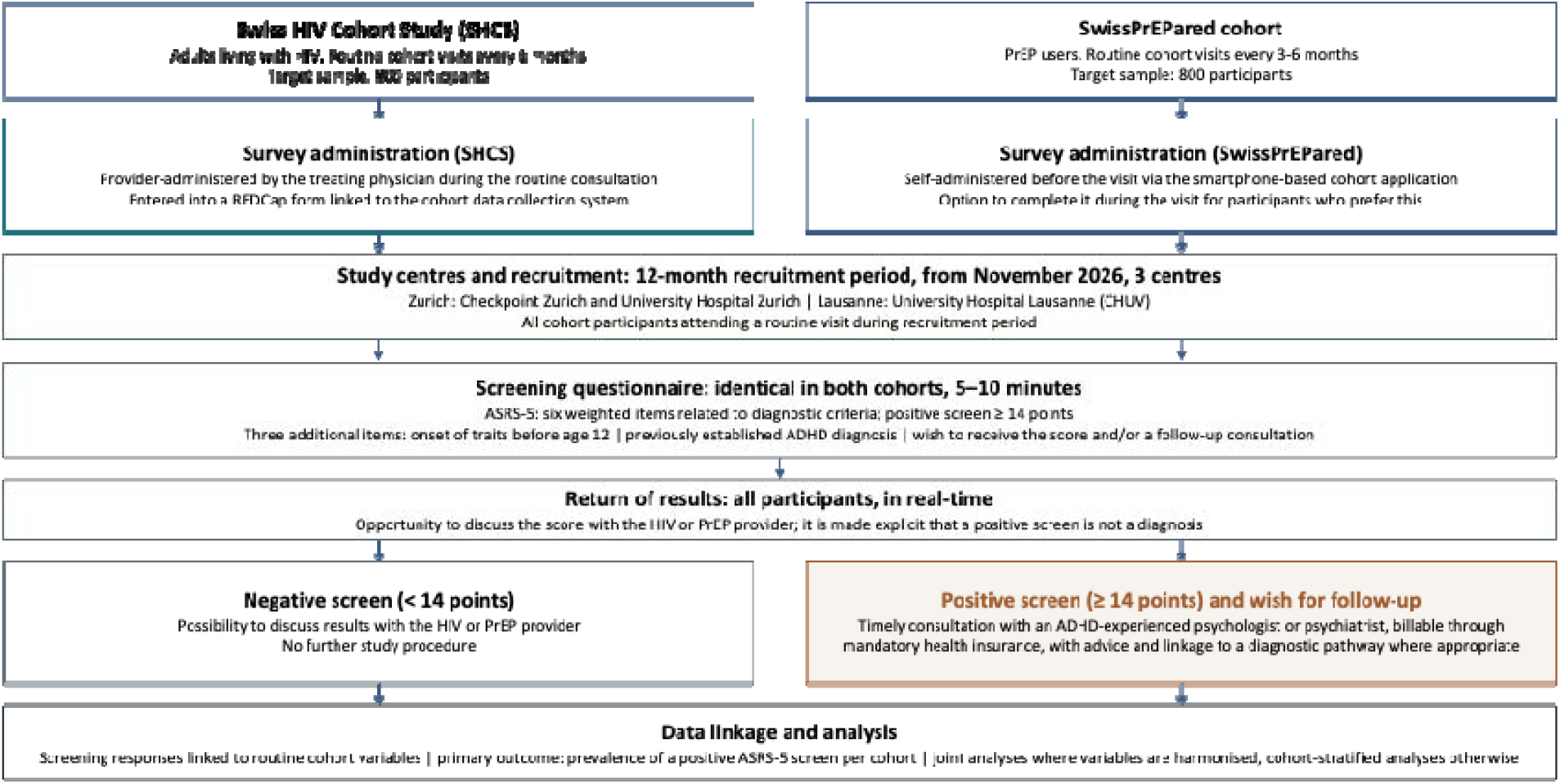
Study flow across the two cohorts and three study centres. Footnote: ADHD, attention-deficit/hyperactivity disorder; ASRS-5, Adult ADHD Self-Report Scale Screener for DSM-5; CHUV, University Hospital Lausanne; PrEP, pre-exposure prophylaxis; SHCS, Swiss HIV Cohort Study; USZ, University Hospital Zurich.

## Measures

### ADHD screening tool: Adult ADHD Self-Report Screening Scale for DSM-5 (ASRS-5)

The ASRS-5 is a widely-used instrument developed by the World Health Organization and the US National Institute of Mental Health to identify traits consistent with adult ADHD.^40,41^ It comprises six items (Part A) drawn from the full 18-item ASRS and is aligned with DSM-5 criteria (Table 1). The items span inattention (item 1), hyperactivity/impulsivity (items 2-4) and executive dysfunction (items 5-6), each rated on a five-point Likert scale referring to traits occurring over the past six months. Items are weighted ^41,42^, and a total score of 14 or higher indicates a positive screen. Against structured clinical interview standards, the ASRS-5 version has reported sensitivity of 91.4%, specificity of 96.0%, an area under the curve of 0.94 and a positive predictive value of 67.3%.^41,43^ In a general population sample, the ASRS-5 screened 11.2% as positive (vs. a true weighted ADHD prevalence of 8.2% in that sample).^41^ The instrument identifies traits and does not establish a diagnosis.

**Table 1.**
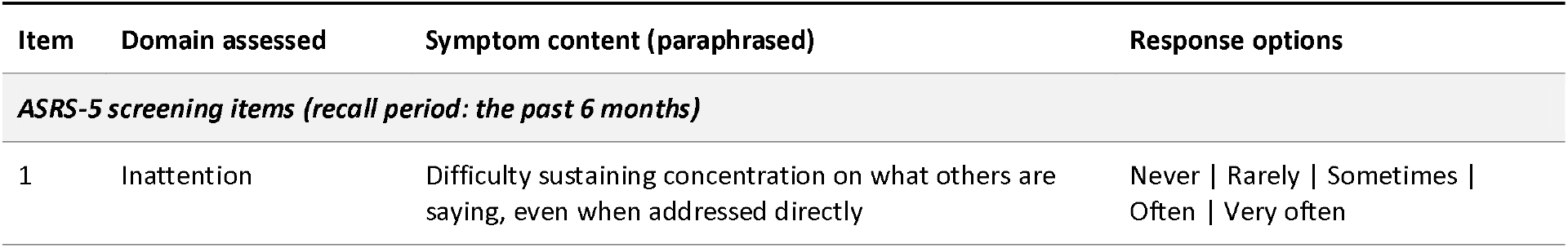
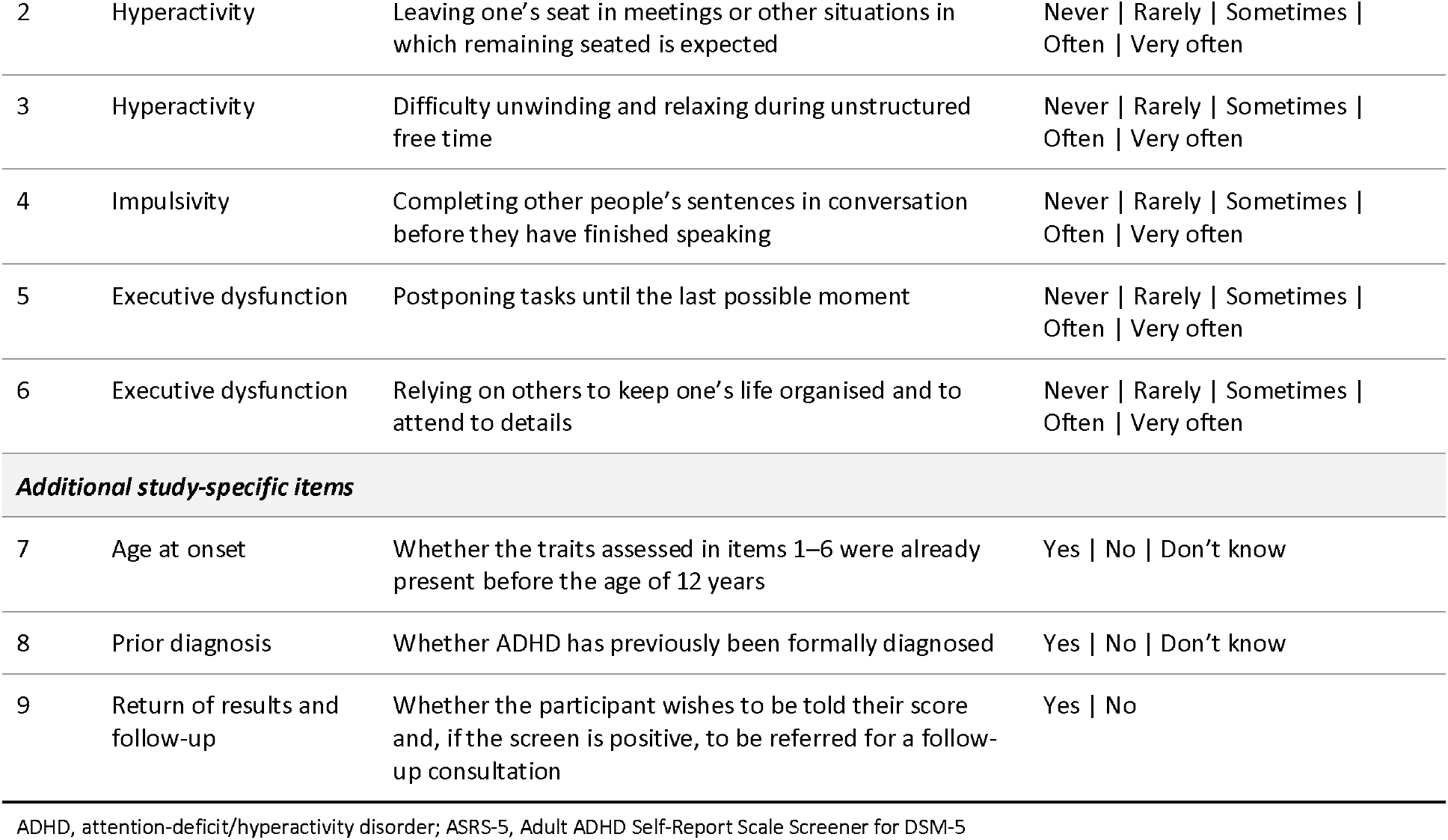
Content and scoring of the six-item Adult ADHD Self-Report Scale Screener for DSM-5 (ASRS-5) and of the three additional study-specific items.

### Additional questions and linked cohort data

Alongside the ASRS-5, three further questions capture (i) whether the assessed traits began before the age of 12, (ii) any previously established ADHD diagnosis, and (iii) whether the participant wishes to learn their score and, in the case of a high score, be referred for follow-up. Screening responses will be linked to routinely collected cohort data, including sociodemographic variables (age, education, employment, co-occurring conditions, sex assigned at birth, gender identity and sexual orientation), substance use, sexual behaviour and STIs, mental health indicators, and treatment, adherence and viral-load data. Because some variables are recorded differently across the two cohorts, an overview of variables and their cross-cohort comparability will inform which analyses are conducted jointly versus within cohorts and will be published with results. A short pilot will test the questionnaire with five SHCS and five SwissPrEPared providers and participants before the recruitment starts.

### Referral pathway and site training

Every participant has the possibility to discuss the result with the health care professional during HIV or PrEP visits. HCPs will be trained in ADHD sensitive care and in providing basic information on ADHD. Participants with a positive score on the ASRS-5 (see Figure 1) will in addition be offered a one-time consultation with a psychiatrist or psychologist experienced in ADHD, to discuss the implications of their screening score, and provide advice and linkage to a diagnostic pathway if appropriate. This consultation is billable through mandatory health insurance. Checkpoint Zurich has an established internal triage and counselling pathway, while CHUV and USZ participants will be offered an equivalent service through independent ADHD specialists. To prepare participating sites for the study and to respond to our aim to raise awareness about ADHD among healthcare professionals, each site will receive training which, beyond covering the study rationale and design, the questionnaire, and the referral pathway, will provide an overview of ADHD and its implications.

### Sample size

The sample size is based on estimating a proportion with specified precision, using n = Z^2^ x p(1−p) / d^2^, with Z = 1.96 (95% confidence) and a margin of error d of ±0.03.^44^ For the SHCS, an expected ADHD-trait prevalence of 20-25%, informed by the two small prior HIV studies,^25,26^ yields a target of 800 participants. For SwissPrEPared, where no prevalence data exist, we anchor on UK Adult Psychiatric Morbidity Survey data (2023/24), in which 13.9% of adults screened positive on the ASRS-5,^45^ and conservatively assume an elevated prevalence of 20-25%, yielding the same target of 800 participants as across the SHCS. Larger samples would increase precision for the secondary objectives, hence, additional participating sites may be added and/or individual participant numbers increased in case of rapid recruitment.

### Data management

Data are entered into the routine data collection systems of each cohort. Regular data checks will be conducted across both data centres, and a dashboard will monitor recruitment. Data handling will comply with SHCS and SwissPrEPared governance policies.

### Statistical analysis

Analyses will be primarily descriptive and exploratory, consistent with the cross-sectional design. All tests will be two-sided at a significance level of 0.05. Given the exploratory nature of the secondary analyses, patterns and confidence intervals will be prioritised over p-values. Analyses will be conducted jointly where variables are harmonised across cohorts, with cohort membership included as a covariate or stratification factor, and separately within each cohort otherwise. ADHD traits are defined as a binary variable (positive ASRS-5 screen, ≥ 14 points).

For the primary objective, prevalence will be calculated as the number of participants with a positive screen divided by the total number of respondents, with 95% Wilson score confidence intervals.^46^ For the secondary objectives, prevalence will be compared between cohorts using a chi-square test, sociodemographic profiles will be characterised through stratified descriptive analyses and unadjusted group comparisons, presented as distributions and absolute prevalence differences with 95% confidence intervals, without multivariable modelling.^47^ Associations between ADHD traits (as exposure) and the specified behavioural, mental health, adherence and treatment modality outcomes will be examined using individual multivariable models per outcome, with a small number of temporally and clinically meaningful adjustment variables, and directed acyclic graphs to justify covariate choice. Gender identity and sexual orientation among PrEP users will be described in the same way as the sociodemographic profiles.

The primary outcome is expected to have minimal missingness owing to embedded questionnaire logic, and no imputation will be performed for the ASRS-5 outcome. Missingness in covariates will be described by variable and cohort, complete-case analysis will be the primary approach. Prespecified sensitivity analyses will (i) include vs exclude participants with a previously recorded ADHD diagnosis and (ii) compare the characteristics of participants who completed the questionnaire with those who did not, using routinely collected cohort data, to assess potential selection bias.

### ETHICS

The study is a nested project within the SHCS and SwissPrEPared and falls under their existing ethical approvals. All participants provided general data collection consent on joining the cohorts. At the start of the survey it will be made clear that the ASRS-5 is a screening tool and not a diagnostic assessment, that individual results will not affect medical care or cohort participation, that follow-up will be proposed to participants with elevated scores, and that any personal data will be handled confidentially in line with cohort standards.

### Dissemination

Findings will be published in peer-reviewed journals, presented at conferences. Results will also be published on the study website (www.adhd-hiv-prep.com) and further returned to participants and communities through lay summaries on the SHCS and SwissPrEPared platforms and through community communication channels. Results will also be the subject of an episode of the SHCS podcast series.

## DISCUSSION

This is the first study to systematically estimate the prevalence of ADHD traits at large scale among adults living with HIV and users of PrEP.^48^ We are aware of only two surveys exploring this question so far, one from India and one from Turkey. They were small scale (n=100; n= 85) and found that between 20-29% of adults living with HIV also lived with ADHD, based on a full psychiatric diagnostic process.^25,26^

Logistical challenges including complex and time-consuming diagnostic procedures, a shortage of trained healthcare professionals, and resultant long waits for appointments, mean that a full diagnostic process is neither desirable nor feasible at the scale of the current study. Indeed, this would constitute an unreasonable burden on both medical staff and participants in the absence of a systematic screening of traits indicating which participants might benefit from advice concerning diagnosis. This informs our approach of initially screening all participants with the ASRS-5 and then linking those with a positive score indicating the interest of pursuing a diagnostic pathway with a specialist. A similar approach was recently taken by a Belgian research team when screening for autism traits amongst people living with HIV and accessing PrEP.^35,49^

Beyond describing prevalence of ADHD traits and behavioural and clinical patterns across the two cohorts, the study is intended to raise awareness and inform HIV and PrEP care. Quantifying the potential burden of previously unrecognised ADHD may support consideration of targeted screening and referral pathways within HIV and PrEP care. The planned analyses regarding treatment adherence^50^, mental health, sexual behaviour and interest in long-acting HIV treatment modalities are exploratory, but could help identify subgroups for whom tailored support, such as long-acting HIV prevention and treatment options that depend less on day-to-day adherence, would be especially valuable. Further, the study may inform future PrEP recommendations. The study is intended to generate hypotheses and to provide a foundation for further planned longer-term research on the topic.

The study’s principal strengths are its originality, its scale, its systematic sampling, its embedding in routine care, and its linkage to rich, prospectively collected data across two large nation-wide cohorts. There are several potential limitations. The ASRS-5 identifies traits, not diagnoses, hence, the study measures screening-positive prevalence, which will exceed the prevalence of diagnosed ADHD, and its positive predictive value depends on an underlying prevalence that is currently uncertain. The cross-sectional design supports description and patterns but not causal inference. Differing administration modes (provider-administered in the SHCS, self-administered in SwissPrEPared) and incomplete harmonisation of some variables constrain some of the between-cohort comparisons. Residual selection bias cannot be excluded despite systematic sampling, which we will address by comparing responders with non-responders.

Screening for stigmatising realities such as HIV, PrEP use, and ADHD carries risks: of compounding stigma^49–51^, of re-enforcing neurodivergent human variation as pathology^31^, and of implying that ADHD traits inherently explain poor sexual health outcomes. We take these risks seriously and have attempted to design the study to resist them. Throughout, we decline to frame ADHD as inherently causing poor sexual health outcomes, and present findings as evidence of unmet needs and structural gaps in care that intersect with and aggravate the impact of traits associated with the condition. Referral after a positive screen is offered and opt-in never imposed. The study is community-driven, with experiential knowledge embedded in the research team and in external review. Results will thus be interpreted with, rather than just about, the people they concern. Informed by Critical ADHD Studies, our aim here is not to present ADHD simplistically as an individual pathology, but to make care more responsive to it and raise awareness amongst HIV healthcare professionals, treating an ADHD positive screen as holding potential to improve support, understanding, and choice.^29,31^

## Data Availability

All data produced in the present work are contained in the manuscript

## DECLARATIONS

### Contributors

DJP, BH and AA conceived the study and are principal investigators. SC provided in-depth technical and scientific input. GHY, MC, IAA, SC and SCO contributed to the study design and reviewed the survey implementation plan. MC, IAA and DJP are part of the SHCS scientific board and BH leads SwissPrEPared. FK, MC, IAA and BH are site investigators. All authors reviewed and approved this protocol.

### Patient and public involvement

see statement in main text

### Members of the Swiss HIV Cohort Study

Abela IA, Aebi-Popp K, Anagnostopoulos A, Bernasconi E, Boyd A, Braun DL, Bucher HC, Calmy A, Cavassini M (Chairman of the Clinical and Laboratory Committee), Chaudron SE (Head of Data Centre), CiuffiA, Dollenmaier G, Egger M, Elzi L, Fehr JS, Fellay J, Frigerio Malossa S, Fux CA, Günthard HF, Hachfeld A, Haerry DHU (deputy of “Positive Council”, patient representative), Hasse B, Hoffmann M, Huber M, Jackson-Perry D, Kahlert CR, Kaufmann D, Keiser O, Kouyos RD, Kovari H, Kusejko K, Labhardt ND, Leuzinger K, Marzolini C, Metzner KJ, Müller N, Nemeth J, Nicca D, Notter J, Paioni P (Chairman of the Mother & Child Substudy), Perreau M, Rauch A (President of the SHCS), Salazar-Vizcaya LP, Schmid P, Segeral O, Speck RF, Stöckle M, Surial B, Tarr PE, Trkola A, Wandeler G (Chairman of the Scientific Board), Weisser M, Yerly S.

### Members of SwissPrEPared

J. Fehr (Scientific Sponsor), B. Hampel (Principal Investigator), A. Farnham (Scientific manager), Bernard Surial, Fanny Joye, Philip Bruggman, Nicola Low, Florian Vock, Julia Notter, Dominique Braun, Claudia Bernardini, Katharina Kusejko, Matthias Hoffmann, Guex Johanne

